# Characteristics and Outcomes of Hospitalized Young Adults with Mild Covid-19

**DOI:** 10.1101/2020.06.02.20106310

**Authors:** Arvind Kumar Sharma, Asrar Ahmed, Vaseem Naheed Baig, Prahalad Dhakad, Gaurav Dalela, Sudhanshu Kacker, Vasim Raja Panwar, Raja Babu Panwar, Rajeev Gupta

## Abstract

**Background:** Stormy course has been reported among hospitalized adults with covid-19 in high- and middle-income countries. To assess clinical outcomes in consecutively hospitalized patients with mild covid-19 in India we performed a study.

**Methods:** We developed a case registry of successive patients admitted with suspected covid-19 infection to our hospital (n=501). Covid-19 was diagnosed using reverse transcriptase polymerase chain reaction (RT-PCR). Demographic, clinical, investigations details and outcomes were recorded. Descriptive statistics are presented.

**Results:** Covid-19 was diagnosed in 234 (46.7%) and data compared with 267 (53.3%) negative controls. Mean age of covid-9 patients was 35.1±16.6y, 59.4% were <40y and 64% men. Symptoms were in less than 10% and comorbidities were in 4-8%. History of BCG vaccination was in 49% cases vs 10% controls. Cases compared to controls had significantly greater white cell (6.96+1.89 vs 6.12+1.69×10^9^ cells/L) and lower lymphocyte count (1.98+0.79 vs 2.32+0.91×10^9^ cells/L). No radiological and electrocardiographic abnormality was observed. All these were isolated or quarantined in the hospital and observed. Covid-19 patients received hydroxychloroquine and azithromycin according to prevalent guidelines. One patient needed oxygen support while hospital course was uncomplicated in the rest. All were discharged alive. Conversion to virus negative status was in 10.2±6.4 days and was significantly lower in age >40y (9.1±5.2) compared to 40-59y (11.3±6.1) and ≥60y (16.4±13.3) (p=0.001).

**Conclusions:** This hospital-based registry shows that mildly symptomatic or asymptomatic young covid-19 patients have excellent prognosis.

What is already known on this subject?
- High rates of morbidity and mortality following hospitalization for covid-19 has been reported from most developed countries.
- In most developing countries covid-19 patients with mild disease are also hospitalized due to lack of home isolation. Clinical outcome of such patients has not been well studied.

What this study adds?
- We performed a clinical registry of successive young patients with mild covid-19 hospitalized with minimal symptoms and comorbidities.
- None of the patients developed complications and all were discharged alive.

## INTRODUCTION

Covid-19 (SARS-CoV-2 infection) that was initially detected towards the end of December 2019 is now a worldwide pandemic.^1^ In the early phase of this pandemic the disease burden was high in East Asian countries but the epicenter shifted rapidly to Europe and presently North America has the highest burden.^2^ Among the larger countries, increasing trend in case burden and deaths is presently being reported from United States, Brazil, Russia, Mexico and India (BRIMUS).^3^ The clinical course of covid-19 ranges from asymptomatic carrier stage to a fulminant course characterized by high grade fever, dyspnea, hypoxia and pulmonary and cardiovascular collapse. Case-registries from China, France, Italy, UK and US have reported that more than 20% patients need oxygen support and about 10-15% need invasive ventilation.^4-13^ Mortality varies from 5-15% in hospitalized patients in various series although population level data suggests a lower mortality varying from 1% to 10%.^14^

Data from publicly available resources in India have reported disease prognosis similar to other countries and adverse outcomes range from 2-10% patients.^15^ Clinical burden of covid-19 has presently shifted to lesser developed states in India and is present in younger population.^15^ The mortality varies from state to state with high numbers reported from Maharashtra and Gujarat. ^15^ A small case-series (n=20) from Rajasthan reported a benign clinical presentation and no fatalities.^16^ Dread of this disease is widespread globally and especially in lower-middle income countries where social media has generated an information epidemic destroying livelihoods.^17,18^ Suicides have been reported among persons even with mild disease.^17^ No previous registries have reported outcomes in low socioeconomic patients with mild disease at admission from India. To describe clinical course and outcomes in successively hospitalized patients from mild covid-19 infection at a university hospital we performed a registry-based study in Rajasthan, India.

## METHODS

We obtained data on successively presenting patients with suspected or confirmed covid-19 at a 530-bed government hospital designated by the state government as covid-19 quarantine, isolation and treatment centre. The study has been approved by the college administration and institutional ethics committee (Registration No. ECR/762/Inst/RJ/2015) was notified accordingly. Ethics committee clearance has been obtained for publication of this report.

All individuals who had history of contact as well as confirmed patients with covid-19 are admitted to various covid-19 designated hospital in the state of Rajasthan (2020 population 77.1 million). We recruited all individuals who were either confirmed covid-19 cases or suspected due to history of contact with patients. Epidemiological, clinical, laboratory, and radiological characteristics, treatment and outcomes data were obtained using history, electronic medical records and transcribed in data collection forms. Diagnosis of covid-19 infection was made using history of contact with a patient or living in high-risk location, fever, cough, pain throat, anosmia, ageusia or dyspnea and confirmed according to SARS-CoV-2 nucleic acid real time polymerase chain reaction (RT-PCR) tests using nasopharyngeal specimens collected using standard protocols.^19^

According to the diagnostic criteria and RT-PCR test results at admission we divided the cohort into two groups: covid-19 positive (n=234) and covid-19 negative (n=267). We isolated covid-19 positive patients in isolation wards or intensive care unit depending on clinical condition while covid-19 negative patients were quarantined on a separate floor. The diagnostic tests were repeated for each patient over a 2-week period. RT-PCR samples for covid-19 were repeated after every 72 hours to determine timing of conversion to non-infective status. The patients were discharged from the hospital when there was clinical improvement along with evidence of clearance of virus as confirmed by two negative RT-PCR test results at least 24 hours apart.

### Statistical analyses

The data were computerized and data processing was performed using commercially available software (SPSS v.20.0). Data are expressed either in numbers ± 1 S.D. or as proportions (percent). Significance of differences in cases vs controls for numerical variables was performed with unpaired t-test and X^*2*^-test was used for nominal variables. P values of <0.05 is considered significant.

## RESULTS

We recruited successive persons hospitalized with suspected covid-19 infections over a period of 6 weeks from 20^th^ March to 30^th^ April 2020 (n=501). There were 234 covid-19 positive cases and 267 covid-19 negative cases, mostly from households of positive patients. Data from this group were analyzed as controls.

Descriptive statistics of the study cohort are in Table 1. Most of the patients were from urban locations and about half of them were from urban slums. Mean age of patients was 35.1±16.6 years with 33.3% in age-group 40-59y and 6.8% in age-group 60+. All the cases and controls were recruited during screening of affected locations in Jaipur, most were asymptomatic, and less than 10% cases and 2% controls had any symptoms. Mean symptoms duration was 4.8±2.0 days in cases and 5.0±2.6 in controls. Co-morbidities such as hypertension, diabetes and COPD were observed in a small minority and smoking was infrequent. History of BCG vaccination was available in half of the cases and 10% controls. Hematological and biochemical investigations were performed in 149 cases and 128 controls. Other patients refused investigations. Cases had significantly higher total white cell counts and lower absolute lymphocyte counts. Transaminases were also greater in cases compared to controls. No significant findings were reported in chest radiographs or electrocardiograms in both the groups (Table 2).

**Table 1:** Sociodemographic and clinical characteristics of covid-19 positive patients and controls.

|  |  | **Covid-19**<br>**cases (n=234)** | **Suspected**<br>**Contacts, controls (n=267)** | **P value**<br>**(t-test or $\chi^2$)** |
| --- | --- | --- | --- | --- |
| Male: female |  | 151:83 | 153:114 | 0.099 |
| Residence | Urban | 132(56.4) | 195(73.0) | <0.001 |
|  | Urban slum | 102(43.6) | 72 (27.0) |  |
|  | Rural | -- | -- |  |
| Age | Mean + SD | 35.1±16.6 | 28.4±17.8 | <0.001 |
|  | Median (IQR) | 35 (12-58) | 25 (10-40) |  |
| Age groups | <20 | 37 (15.8) | 90 (33.4) | <0.001 |
|  | 20-39 | 102 (43.6) | 108 (40.5) |  |
|  | 40-59 | 78 (33.3) | 49 (18.4) |  |
|  | 60+ | 16 (6.8) | 19 (7.1) |  |
| Symptoms | Fever | 10 (4.3) | 5 (1.9) | 0.116 |
|  | Cough | 12 (5.1) | 5 (1.9) | 0.045 |
|  | Taste/smell disorder | -- | -- | 0 |
|  | Breathlessness | 10 (4.3) | 2 (0.8) | 0.010 |
|  | Others | 3 (1.3) | 0 (0) | 0.063 |
| Symptom duration (days) | Mean | 4.8±2.0 | 5.0±2.6 | 0.815 |
|  | Median (IQR) | 5 (2-8) | 3 (0-7) |  |
| Co-morbidities | Hypertension | 11 (4.7) | 2 (0.8) | .006 |
|  | Diabetes | 11 (4.7) | 4(1.5) | .036 |
|  | COPD/Asthma | 12(5.1) | 3 (1.1) | 0.015 |
|  | Coronary heart disease | -- | -- | -- |
|  | Smoking | 20(8.5) | 1(0.4) | <0.001 |
|  | Others | -- | -- | -- |
| BCG Vaccination | Yes | 115(49.2) | 26(9.7) | <0.001 |
|  | No | 112(47.9) | 237(88.8) |  |
|  | Don't know | 7(3.0) | 4(1.5) |  |
| Physical exam | BP (systolic) | 119.1±11.1 | 116.7±11.1 | 0.016 |
|  | BP [diastolic] | 78.5±8.3 | 76.8±6.4 | 0.013 |
|  | Chest crepts | -- | -- | -- |
| Drug therapy | Anti-pyretic | 10 (4.3) | 5 (1.9) | 0.116 |
|  | Cough suppressants | 12 (5.1) | 5 (1.9) | 0.045 |
|  | Hydroxychloroquine | 234(100) | -- | -- |
|  | Azithromycin | 234(100) | -- | -- |
|  | Other antibiotics | -- | -- | -- |
| Hospital duration | <10 days | 33(14.1) | 84(31.5) | <0.001 |
|  | 10-14 days | 56(23.9) | 152(56.9) |  |
|  | 15-19 days | 37(15.8) | 31(11.6) |  |
|  | 20 days+ | 108(46.1) | -- |  |
|  | Median duration (IQR) | 18 (5-31) | 12 (8-16) | <0.0001 |
| IQR inter quartile range |  |  |  |  |

**Table 2:** Hematological, biochemical and other investigations.

|  |  | **Covid-19**<br>**Cases (n=149)** | **Suspected**<br>**Contacts, controls**<br>**(n=128)** | **P value**<br>**(t-test)** |
| --- | --- | --- | --- | --- |
| Hematology parameters | White cell count ($10^9$ cells/L) | 6.96±1.89 | 6.12±1.69 | <0.001 |
| | Neutrophil count ($10^9$ cells/L) | 3.82±1.37 | 3.69±1.09 | 0.421 |
| | Lymphocyte count ($10^9$ cells/L) | 1.98±0.79 | 2.32±0.91 | 0.001 |
|  | Lymphocyte:neutrophil ratio | 0.68±0.14 | 0.79±0.19 | <0.001 |
|  | Hemoglobin | 12.85±1.7 | 13.44±0.84 | <0.001 |
| Biochemical parameters | Creatinine | 0.98±0.42 | 1.01±0.14 | 0.440 |
|  | SGOT (U/L) | 26.9±10.0 | 21.3±5.4 | <0.001 |
|  | SGPT (U/L) | 27.2±15.1 | 22.4±0.5 | 0.001 |
| Chest radiology |  | No significant finding | No significant finding | -- |
| Electrocardiogram<br>(baseline) | General report<br>QTc interval | Normal limits<br>Normal | Normal limits<br>Normal | -- |

All covid-19 patients were prescribed hydroxychloroquine (400mg once daily) and azithromycin (500mg once daily) for 7 days according to the prevalent local as well as guidelines by government of India.^20^. One patient required oxygen support although radiogram of chest was normal. No other complication related to covid-19 was found in any of these patients. There were no deaths in the hospital. Conversion to virus negative status occurred in 10.2±6.4 days (median 8 days, interquartile range 2-14) and was significantly lower in age group >40y (9.1±5.2 days) compared to 40-59 years (11.3±6.1 days) and ≥60 years (16.4±13.3) (p=0.001). All the patients recovered and were discharged from the hospital at a median of 19 days in covid-19 positive group and 12 days in the control group.

## DISCUSSION

This case series of asymptomatic and mildly symptomatic young patients in India shows extremely low rates of covid-19 complications. Only 21 patients (9%) were symptomatic at the time of admission and clinical, biochemical, hematological and radiological abnormalities, reported from larger series elsewhere,^10-13^ were largely absent. Younger age and low incidence of complications is most likely related to the fact that these patients were identified during a house-to-house survey in Jaipur and mostly were asymptomatic carriers and also because only less sick patients are hospitalized with us. More sick covid-19 patients are treated at another government-designated covid-19 centre in the city as reported earlier.^16^ Even in the more sick cohort of older patients with multiple co-morbidities at the second hospital no mortality has been reported. ^16^ Recent data from the state shows that mortality from covid-19 is significantly lower in Rajasthan than other states in India.^15^ Only a limited number of studies have reported a higher rate of asymptomatic patients at the time of hospital admission and a high frequency of asymptomatic patients in this study can be justified by local policies that are guided by poor quality of housing, joint-family structure and limited living space of most of our patients. This makes effective home-quarantining impossible. Similar situation (housing, family structure, etc.) exists in most of the less developed countries.^21-^

Large clinical registries from China, Europe, UK and USA have reported low morbidity and mortality rates in younger patients with covid-19 similar to our experience. A report from the Chinese Centre for Disease Control and Prevention identified 44672 covid-19 patients from 72314 case records of suspected infections.^5^ Most of the patients were in age group 30-79 years with mild disease in 81%. Case fatality rate was 2.3% overall, 8.0% in patients 70-79 and 14.8% in 80+ years, with no deaths reported in mild cases similar to our study. Lechien et at reported initial experience of 1420 patients in Europe from France, Belgium, Spain and Switzerland.^9^ The mean age was 39±12 years, 94% were less than 60 years of age and less than 10% required hospitalizations. In this cohort more than 70% patients were symptomatic, younger had greater ear, nose and throat symptoms while elderly had more generalized symptoms including fever. No mortality was reported in this cohort, similar to our outcomes. In the OpenSAFELY Collaborative report, Williamson et al identified covid-19 risk factors in a population of more than 17 million adults and 5683 patients from February to April 2020. Compared to age group 50-59y, age-sex adjusted hazard ratio (95% confidence intervals) for covid-19 deaths at age 18-39years was 0.05 (0.04-0.08) and 40-49 years was 0.27 (0.21-0.34). Older age and multimorbidity were associated with significantly greater risk.^10^ Richardson et al reported presenting characteristics, comorbidities and outcomes among 5700 patients hospitalized with covid-10 in New York.^13^ The median age was 63y (IQR 52-75y). In the age-groups 20-29, 30-39 and 40-49 years, respectively, the mortality was 7.1%, 4.6% and 8.2% in men and 1.8%, 2.5% and 2.5% in women which is greater than in the present study. Age-specific data on comorbidities and clinical parameters were not reported. On the other hand, studies from Iran, Italy France and Europe have reported significantly higher mortality in hospitalized covid-19 patients.^7,8,10,12^

Apart from younger age and low prevalence of comorbidities in our patients other virus and host related factors could be possible reasons for low complication rates. These include lower viral load,^22^ attenuation of virus pathogenicity,^23^ host genetics,^24^ presence of attenuating antibodies,^25^ rate of development of neutralizing antibodies,^interaction of virus with throat or gut microbiome,^26^ and low or absent cytokine response.^25,27^ Other reasons that have been speculated are high previous exposure to viral and bacterial infections, childhood BCG vaccination and presence of competing infective comorbidities.^28-30^ Only half of our patients had history of BCG vaccination compared to 10% in controls. However, history is poor guide to vaccination in India and this may not be reliable in this low socioeconomic status cohort. South Asian ethnicity has been reported as a risk factor for greater covid-19 disease and deaths in the UK and it has been suggested that both basic and public health research is needed to identify causal pathways of the increased risks and their management.^31^ Every patient in our study received drug treatment with hydroxychloroquine and azithromycin. Although recent trials and revised government of India guidelines^20^ and other recommendations do not support their routine use,^32^ we followed the extant practice. Moreover, use of these drugs has shown to have a neutral effect on outcomes in recent clinical trials.^33^ We do not believe that this is the cause of low morbidity in our cohort and recommend randomized controlled studies for evaluation of efficacy of these drugs.

A major limitation of our study is recruitment of younger patients who are either asymptomatic carriers or those with minimal disease. There is need to study older patients with multiple comorbidities. A small sample size is also a limitation although we included all successive patients admitted to our hospital. This is, also, one of the larger covid-19 registries from India on patients with mild disease. We also did not perform investigations in the whole cohort as many patients refused consent. Chest computed tomographic scan is superior to diagnose early evidence of pulmonary involvement, but due to lack of symptoms and signs of major respiratory involvement and absence of prolonged hypoxia in any of the patients, this was deferred. Furthermore, the duration of RT-PCR positivity from the time of diagnosis in asymptomatic patients may be underestimated due to lack of major symptoms, making it challenging to determine when these cases were infected and become non-contagious.

In conclusion, despite its small sample size, this study is the first and largest report of patients with covid-19 from this region of the country. We have shown that younger patients with low comorbidities have extremely low rate of complications of the disease. This is an important message for the public and policy makers to highlight the essentially benign nature of this disease in the young.

## Data Availability

All the data are reported in the article.

